# Implementation of a Non-Invasive Helmet Ventilation Solution for the Management of Severe COVID-19 Respiratory Disease in Nigeria: The CircumVent Project

**DOI:** 10.1101/2022.08.30.22279372

**Authors:** Aima A. Ahonkhai, Aliyu Abdu, Olukemi Adekanmbi, Nnennaya A. Ajayi, Samuel Ajayi, Happy Akpobi, Ejiro Benjamin Akpochafo, Muktar H. Aliyu, Adaeze C Ayuk, Adedamola A. Dada, Oliver C. Ezechi, Catherine O. Falade, Alex Horstein, Idowu Olusola, Ifeoma Idigbe, Sunday Mogaj, Aleem A. Morenikeji, Baba M. Musa, Nnamdi I. Nwosu, Adenike A Odewabi, Igho Ofokotun, Gbenga Ogedegbe, Onome Ogueh, Temitope O Oyewole, Adeshola I. Sotannde, Alan B. Steinbach, Ifeoma I. Ulasi, Kingsley N. Ukwaj, Uchechukwu S. Unigwe, Olagoke A. Usman, Cyril Uzoke, Adesola Z. Musa, Muyiwa K. Rotimi, Iorhen E. Akase, Wasiu L. Adeyemo, André A. Fenton, Babatunde L. Salako

**Affiliations:** Department of Medicine, Vanderbilt University Medical Center, Nashville, TN, USA; Vanderbilt Institute for Global Health, Vanderbilt University Medical Center, Nashville, TN, USA; Department of Medicine, Bayero University Kano, Kano, Nigeria; Africa Center of Excellence in Population Health and Policy, Aminu Kano Teaching Hospital, Kano, Nigeria; University College Hospital, Ibadan; University of Nigeria Teaching, Oyo, Nigeria; Department of Internal Medicine, College of Health Sciences, Ebonyi State University, Abakaliki/ Alex Ekwueme Federal University Teaching Hospital, Abakaliki, Ebonyi State, Nigeria; Delta State University Teaching Hospital, Oghara, Nigeria; College of Medicine, University of Nigeria / University of Nigeria Teaching Hospital, Ituku-Ozalla, Enugu, Enugu State, Nigeria; Federal Medical Centre, Ebute Metta, Lagos, Nigeria; Nigerian Institute of Medical Research, Lagos, Nigeria; Neurobiology of Cognition Laboratory, Center for Neural Science, New York University, New York, USA; Neuroscience Institute, NYU Langone Medical Center, New York, USA; Inventilator LLC, Providence, RI, USA; Federal Medical Centre, Abeokuta; Ogun, Nigeria; Division of Infectious Diseases, Department of Medicine, Emory University, Atlanta, GA, USA; Institute for Excellence in Health Equity, NYU Grossman School of Medicine, New York, NY, USA; Joint Medical Program, UCB/UCSF, Berkeley, CA; Lagos University Teaching Hospital; Lagos, Nigeria

**Keywords:** SARS-CoV-2, COVID-19 infection, Nigeria, Non-invasive ventilation, Implementation science

## Abstract

Affordable novel strategies are needed to treat COVID-19 cases complicated by respiratory compromise in resource limited settings. We report a mixed-methods pre-post assessment of 1) the useability of CPAP/O2 helmet non-invasive ventilation (NIV) to treat COVID-19, at ∼ 1% the cost of mechanical ventilation; 2) the effectiveness of a train-the-trainer practice facilitation intervention; and 3) whether use of CPAP/O2 helmet NIV was associated with increased COVID-19 infection among healthcare workers. At baseline, eight COVID-19 treatment centers in Nigeria (CircumVent network) received CPAP/O2 helmet systems, and were instructed on its use. After five months, clinicians within the CircumVent netwok participated in a 2-day train-the-trainers educational intervention. The physicians completed i) standardized forms on patient demographics, clinical course, and outcomes for patients seen in the treatment centers; ii) standardized surveys of feasibility and acceptability of use of CPAP/O2 helmet systems; and iii) in-depth-interviews to explore facilitators and barriers to implementation of CPAP/O2 helmet NIV. Physicians described the CPAP/O2 helmet ventilator as easy to use and they felt comfortable training their staff on its use. They rated CPAP/O2 helmet NIV as feasible, acceptable, and appropriate (mean score of 4.0, 3.8, and 3.9 out of 5, respectively, on standardized scales). Case report forms for 546 patients with suspected and/or confirmed COVID-19 infection were obtained between May 2020 and November 2021. Of these, 69% (n=376) were treated before the training; and 29.7% (n=162) were treated with CPAP/O2 helmet ventilation. CPAP/O2 helmet NIV was well-tolerated by patients, with 12% reporting claustrophobia, and 2% reporting loose- or tight-fitting helmets. Although patient outcomes improved among CPAP/O2 helmet users overall, this was not associated with training (P=0.2). This finding persisted after adjustment for disease severity at presentation. Serosurvey of 282 health workers across treatment centers revealed that 40% (n=112) were seropositive for SARS-CoV-2. Seropositivity was significantly associated with direct contact with COVID-19 patients and limited access to PPE and hand hygiene during aerosol generating procedures (P = 0.02), but not use of CPAP/O2 helmet (P’s ≥ 0.2). In conclusion, physicians effectively used CPAP/O2 helmet NIV systems to treat COVID-19 patients in Nigeria without need for practice facilliation of their training and without increased risk of infection among healthcare workers. The use of CPAP/O2 helmet NIV could be an important strategy for treating individuals with COVID-19 infection and other disease conditions complicated by respiratory distress, particularly in settings were resources such mechanical ventilation are limited.

## INTRODUCTION

The coronavirus disease-2019 (COVID-19) pandemic caused by the severe acute respiratory syndrome coronavirus 2 (SARS-CoV-2) is the most devastating infectious disease outbreak in recent times. A common life-threatening complication of COVID-19 disease is hypoxemic respiratory failure often requiring supplemental oxygen therapy, intensive unit care (ICU), and/or mechanical ventilation – interventions that are limited in several low- and middle-income countries (LMICs) (1-4).

To address respiratory compromise in individuals with COVID-19, our multidisciplinary team of clinicians, scientists, entrepreneur, and engineer volunteers (the CircumVent Project) developed and implemented a continuous positive airway pressure and oxygen helmet (CPAP/O2 helmet) non-invasive ventilation (NIV) strategy to reduce the need for invasive ventilation in LMICs. With less than one ventilator per million citizens in Nigeria, invasive mechanical ventilation is a limited therapeutic option in Nigeria and many other LMICs.

Advantages of CPAP/O2 helmet NIV include 1) using low-cost, widely available CPAP devices (1% the cost of a mechanical ventilator); 2) limiting viral dispersion, including potential aerosolization of SARS-CoV-2 by helmet-based filtration; 3) delivering oxygen effectively in part due to positive end-expiratory pressure (PEEP) at relatively low oxygen flow rates, (compared to high oxygen flow using nasal cannula and other forms of NIV), which addresses oxygen scarcity (1, 5-10). Additionally, while data are limited, CPAP/O2 helmet ventilation may have advantages over high flow nasal oxygen, and avoid the need for scarce mechanical ventilation (1, 9, 11). With a population of over 200 million, Nigeria is ideal for assessing this approach (12). Nigeria reported the first case of COVID-19 on February 27, 2020, and has seen a total of 256,000 confirmed cases and 3,143 deaths from COVID-19 disease as of May 31, 2022, a likely under-estimate (13-15). The objectives of this study are to assess: 1) the useability of CPAP/O2 helmet NIV to treat severe COVID-19 infection cases with respitatory distress; 2) the effectiveness of a train-the-trainer practice facilitation intervention; and 3) whether use of CPAP/O2 helmet NIV was associated with increased COVID-19 infection among healthcare workers.

## METHODS

This study was conducted at eight COVID-19 Treatment Centers in Nigeria, “The CircumVent Network,” (Figure 1). A total of 156 CPAP/O2 helmet ventilator kits were delivered to the treatment centers. The CPAP/O2 helmet kits included 22-mm tubing and viral filters for the patient circuit with valves for supplemental oxygen (8). The patient interface is a low-cost subsalve oxygen helmet used in combination with disposable inspiratory and expiratory 22-mm filters, intended to protect healthcare workers from aerosolized viral exposure. The tubing and helmet can be sterilized in a number of ways, including using low-cost disinfectants against SARS-CoV-2 [https://cfpub.epa.gov/wizards/disinfectants/]. All patients who presented to the treatment facilities between May 3, 2020 and November 28, 2021 were eligible for the study, including those that were patients prior to delivery of the CPAP/O2 helmets. Patients were treated with CPAP/O2 helmet based on disease severity and protocol (www.circumventproject.com/protocol). We used a mixed-methods design (8) to address the three study objectives; and utilized the Integrated Promoting Action on Research Implementation Services (iPARIHS) framework to guide program implementation (16). In order to assure buy-in from the treatment centers and Nigeria’s Federal Ministry of Health, we convened a steering committee of key stakeholders across the health sector to provide guidance on implementation strategy.

**Figure 1.**
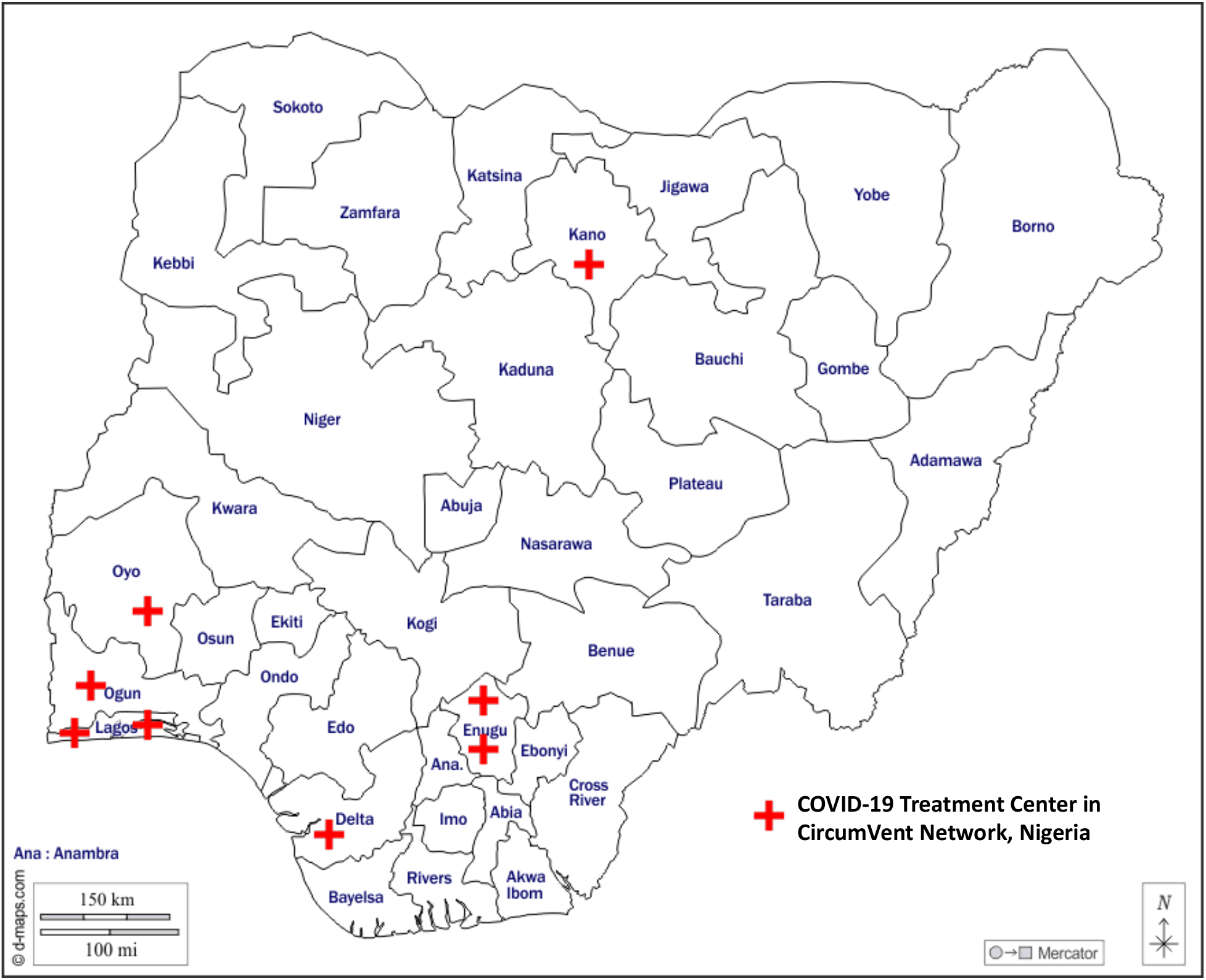
Each red cross represents one of the eight COVID-19 treatment centers in the CircumVent network. All of the centers are housed in tertiary hospitals: Aminu Kano Teaching Hospital (AKTH; 700-bed facility), Delta State University Teaching Hospital (DELSUTH; 250-bed facility), Federal Medical Centre, Abeokuta and Federal Medical Centre Ebute Metta, Lagos (FMC Lagos; 240-bed facility), Lagos University Teaching Hospital (LUTH; 761-bed facility), University College Hospital, Ibadan (UCH; 1000-bed facility), University of Nigeria Teaching Hospital (UNTH; 500-bed facility), Enugu State University Teaching Hospital (EUTH; 50-bed facility), Alex Ekwueme Federal University Teaching Hospital (FUTH; 602-bed facility).

For objective #1, we assessed the feasibility and acceptability of CPAP/O2 helmet use using qualitative interviews and surveys. For usability, we conducted in-depth-interviews with clinicians at the treatment centers. Feasibility, acceptability and appropriateness were determined via in-depth-interviews and assessment via standardized questionnaires, namely Weiner’s feasibility, acceptability and appropriateness of intervention measures (providers persepctives); structured case report forms were used to assess the fit/comfort of the CPAP/O2 helmet devices (patients perspectives) and any oxygen supply issues for each use (17).

For objective #2, a total of 160 clinicians were trained to use CPAP/O2 helmet NIV, and in appropriate clinical indications for use according to our protocol (www.circumventproject.com/protocol). Specifically, patients were eligible for CPAP/O2 helmet NIV treatment if they needed escalation of O2 administration via nasal cannula (1-6 liters per minute), venturi mask (up to 12 liters per minute), or high flow nasal cannula (if available, up to 60 liters per minute) to maintain clinical targets (SPO2 92%, PaO2 >55, resolution of cyanosis, respiratory rate <24 breaths per minute, and Richmond Agitation-Sedation Scale score between −1 and 0 or Glascow Coma Scale [GCS] score ≥11). Patients who were at high risk for aspiration, had decreased mental status (GCS<8), or hemodynamic instability requiring vasoactive agents) were ineligible for helmet use. Patient treatment outcomes assessed at the first of 4 weeks or discharge were compared before and after receipt of the formal training (16).

We compared severity of baseline respiratory illness (respiratory rate and oxygen saturation) among patients who were managed with versus without CPAP/O2 helmet NIV. Clinical outcomes were and included improvement in clinical status (assessed by the treating physician), intubation or death. All treatment centers documented baseline clinical and demographic characteristics, along with clinical outcomes using structured data collection forms in REDCap (18). The characteristics and outcomes were determined as percentages of the patients before and after practice facilitation training with and without CPAP/O2 helmet treatment. Binomial tests of proportions or Chi-squared tests were used for comparisons, as appropriate.

For objective #3, we conducted a retrospective and nested case-control study with a convenience sample of clinical providers who provided clinical care within the CircumVent network. We used a standardized tool to collect information on healthcare and community-related COVID-19 exposures. We collected serum from consenting providers, and assessed for antibodies to SARS-CoV-2 (Table 3), before COVID-19 vaccination was available for healthcare providers in Nigeria. We then compared the working conditions and infectious exposures of the seropositive (cases) and seronegative (controls) individuals.

### Ethics Statement

Ethical approval was obtained from the National Health Research Ethics Committee of Nigeria (protocol number NHREC/01/01/2007). The study was determined to have an exempt status because it is a quality improvement study (Fig. 2). Consequently, formal consent was not obtained. Evaluation of SARS-CoV-2 infection prevalence amongst healthcare workers was conducted with written consent under a National Health Research Ethics Committee of Nigeria protocol.

**Figure 2.**
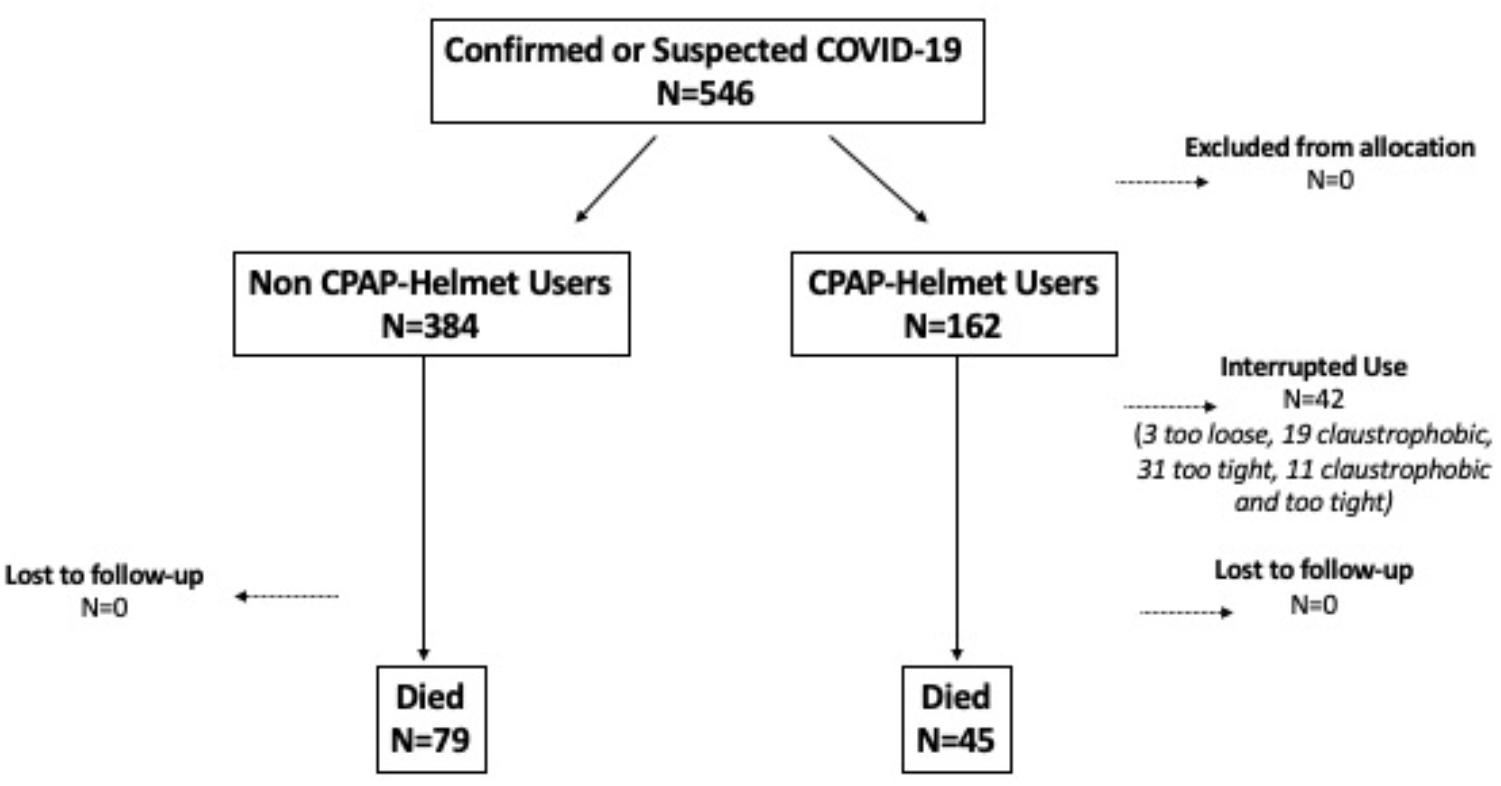
CONSORT flow diagram for the CircumVent Project

### Trial Registration

This trial was retrospectively registered (NCT04929691) with clinicaltrials.gov on June 18, 2021.

## RESULTS

### Feasibility and acceptability of CPAP/02 Helmet NIV (Provider Perspective)

Feasibility, acceptability and appropriateness of the use of the CPAP/O2 helmet was assessed with the Feasibility of Intervention Measure (FIM), Acceptability of Intervention Measure (AIM), and Intervention Appropriateness Measure (IAM) scales (17). A total of 38 clinicians from the eight treatment centers completed the surveys. Average FIM, AIM, and IAM scores were 4.0 (SD 1.0), 3.8 (SD 1.2), and 3.9 (SD 1.1) respectively on a 5-point ordinal scale, with higher scores indicating higher levels of acceptability, feasibility and appropriateness.

Usability was assessed via in-depth qualitative interviews with 16 clinicians (average of 16 years of clinical experience). While most had used CPAP or BIPAP modes of NIV in their practice, the O2 helmet unit was new to all of them. They felt the helmet was an easy-to-use, and innovative mechanism that their teams could deploy with effective training. They stated that adoption and sustainable use of the CPAP/O2 helmet NIV would depend on several factors, including: 1) availability of a constant source of back-up power for usage; 2) availability of consumables and supplies to ensure high flow of oxygen required for the unit (i.e., PEEP valve, O2 bleed adaptors, CPAP hose); 3) provision of helmets with different sizes; and 4) identification/involvement of key administrative stakeholders, including hospital leadership, department chairs, and senior physicians to ensure buy-in for sustainability and maintenance of device use.

Based on findings from the in-depth-interviews, the treatment protocol and training were refined by: 1) modifying the training activities and assigning step-down training responsibility to designated trainers at each COVID-19 treatment center; 2) investigating options for a battery-supplied function; 3) identifying a multi-tank strategy and patient circuit for increasing oxygen flow beyond the 15 L/min limit set by locally available oxygen flow meters; 4) supplying additional helmet sizes and retraining on adjusting the gasket neck size; and 5) involving key administrative stakeholders, specifically an administratively senior individual was formally recruited to accept the responsibilities of principal investigator at each implementation site for organizing and reporting implementation practices (8).

### Clinical Cohort Description

Treating providers in the CircumVent network documented 546 cases of confirmed or suspected COVID-19 infection (between May 3, 2020 and November 28, 2021; 69% of these cases were treated before the training on CPAP/O2 helment use. Mean age of the patient cohort was 52 years, and 62% were men. Hypertension was the most common comorbidity (50%), followed by heart disease (27%), diabetes (24%), and lung disease (21%). Thirty percent of of the 546 patients were treated with CPAP/O2 helmet NIV while 70% were treated according to local standard of care. Comorbidity was more prevalent among CPAP/O2 helmet treated patients compared to those treated via standard of care (hypertension 55% vs 40%, heart disease 31% vs, 19%, diabetes 23% vs. 20%, lung disease 35% vs. 10% respectively). CPAP/helmet-treated patients were more likely to have had baseline tachypnea (respiratory rate ≥20 breaths per minute; 100% vs. 96%, p=0.015), or severe hypoxemia (pulse oximetry <88% than patients treated according to standard of care.

The CPAP/O2 helmet was tolerated by most of the 162 patients (Table 2); 19% had interruptions in CPAP/O2 helmet use due to helmet size or claustrophobia. Twelve percent of patients reported some claustrophobia, 1% felt the helmet was too loose, and 23% reported the helmet was too tight. Inadequate oxygen supply was reported infrequently overall (6% of patients), but more commonly among patients treated with CPAP/O2 helmet compared to those without (11% vs. 4%, p<0.01). Overall, 65% of patients improved (at the first of 4 weeks or discharge), 23% died, and clinical outcomes were missing or unknown for 9% of patients,

### Comparison of clinical features and outcomes among CPAP/02 helmet NIV treated patients pre- and post-training on CPAP/02 helmet NIV

Among the 162 patients treaated with CPAP/02 helmet NIV, 46% (n=74) were treated before and 54% (n=88) were treated after practice facilitation. After practice facilitation, average age of treated patients increased, and prevalence of all of the most common comorbidities increased (hypertension, heart disease, diabetes) except for lung disease [Table 1]. The prevalence of moderate to severe respiratory distress (RR>19) and the prevalence of moderate to severe hypoxia (<90%) were not significantly different in patients treated before or after practice facilitation [Table 2].

**Table 1.** Baseline demographics

|  | **Patients without CPAP/O2 Helmet**<br>**N=384** |  | **Patients with CPAP/O2 Helmet**<br>**N=162** |  | **All Patients**<br>**N=546** |  |
| --- | --- | --- | --- | --- | --- | --- |
|  | **Before Training**<br>**N=302** | **After Training**<br>**N=82** | **Before Training**<br>**N=74** | **After Training**<br>**N=88** |  |  |
| **Sex (n, %)** |  |  |  |  |  |  |
| Male | 190, 63.3% | 52, 63.4% | 45, 62.5% | 47, 53.4% | 334 | 61.6% |
| **Age (yrs) avg. ± SD** | 52 ± 17.8 |  |  |  |  |  |
| <25 | 22, 7.3% | 2, 2.4% | 6, 8.1% | 8, 9.1% | 38 | 7.0% |
| 25-34 | 30, 9.9% | 6, 7.3% | 8, 10.8% | 0, 0.0% | 44 | 8.1% |
| 35-44 | 54, 17.9% | 10, 12.2% | 15, 20.3% | 9, 10.2% | 88 | 16.1% |
| 45-54 | 57, 18.9% | 12, 14.6% | 7, 9.5% | 22, 25.0% | 98 | 17.9% |
| ≥55 | 124, 41.1% | 51, 62.6% | 36, 48.6% | 49, 55.7% | 260 | 47.6% |
| Missing | 15, 5.0% | 1, 1.2% | 2, 2.7% | 0, 0.0% | 18 | 3.3% |
| **Marital Status (n, %)** |  |  |  |  |  |  |
| Married | 211, 81.6% | 70, 85.4% | 59, 81.9% | 72, 81.8% | 412 | 82.4% |
| Single | 32, 12.4% | 9, 11.0% | 9, 12.5% | 6, 6.8% | 56 | 11.2% |
| Widowed | 15, 5.8% | 3, 3.7% | 4, 5.6% | 10, 11.4% | 32 | 6.4% |
| **Education (n, %)** |  |  |  |  |  |  |
| No formal education | 6, 2.5% | 2, 2.5% | 3, 4.5% | 2, 2.3% | 13 | 2.8% |
| Primary | 20, 8.5% | 4, 4.9% | 2, 3.0% | 4, 4.6% | 30 | 6.4% |
| Secondary | 79, 33.5% | 23, 28.4% | 12, 17.9% | 24, 27.6% | 138 | 29.3% |
| Tertiary | 131, 55.5% | 52, 64.2% | 50, 74.6% | 57, 65.5% | 290 | 61.6% |
| **Comorbidity (n, %)** |  |  |  |  |  |  |
| Lung Disease | 23, 10.3% | 15, 18.3% | 31, 49.2% | 25, 28.7% | 94 | 20.7% |
| Hypertension | 104, 41.1% | 49, 59.8% | 35, 53.0% | 54, 61.4% | 242 | 49.5% |
| Heart Disease | 37, 16.7% | 35, 42.7% | 16, 25.0% | 35, 39.8% | 123 | 27.0% |
| Diabetes | 55, 22.6% | 22, 26.8% | 11, 16.7% | 27, 30.7% | 115 | 24.1% |
| Immunosuppression | 16, 7.2% | 19, 23.2% | 5, 7.8% | 7, 8.0% | 47 | 10.3% |
| Cancer (past 12 mos) | 4, 1.8% | 4, 4.9% | 2, 3.2% | 3, 3.4% | 13 | 2.9% |
| Pregnant | 7, 3.3% | 2, 2.5% | 5, 8.2% | 1, 1.2% | 15 | 3.4% |

**Table 2:** Clinical characteristics of covid-19 patients in multi-site Nigerian cohort treated with and without CPAP/O2 helmet NIV, before and after the training. (*breaths per minute).

|  | Patients without CPAP/O2 Helmet<br>N=384 |  | Patients with CPAP/O2 Helmet<br>N=162 |  | All Patients<br>N=546 | P Value |
| --- | --- | --- | --- | --- | --- | --- |
|  | Before Training<br>N=302 | After Training<br>N=82 | Before Training<br>N=74 | After Training<br>N=88 |  |  |
| **Respiratory Rate\* Classification (n, %)**<br>11-19, Mild<br>20-24 Moderate<br>25-30 Severe<br>>30 or <10 Very Severe | 11,3.7%<br>85,28.3%<br>73,24.3%<br>131,43.7% | 4, 4.9%<br>14, 17.1%<br>27, 32.9%<br>37, 45.1% | 0, 0.0%<br>7, 9.7%<br>22, 30.6%<br>43, 59.7% | 0, 0.0%<br>25, 28.4%<br>18, 20.8%<br>45, 51.1% | 15, 2.7%<br>131, 24.0%<br>140, 25.6%<br>256, 46.9% | **Before**<br>**P=0.02** |
|  |  |  |  |  |  | After<br>P=0.24 |
| **Pulse Oximetry (n, %)**<br>≥ 90, Mild<br>< 90, Moderate<br>< 88, Severe | 170, 58.8%<br>27, 9.3%<br>92, 31.8% | 42, 51.2%<br>9, 11.0%<br>31, 37.8% | 28, 37.8%<br>15, 20.3%<br>31, 41.9% | 35, 40.2%<br>11, 12.6%<br>41, 47.1% | 275, 50.4%<br>62, 11.4%<br>195, 35.7% | **Before**<br>**P=0.002** |
|  |  |  |  |  |  | After<br>P=0.354 |
| **Inadequate O2 (n, %)**<br>Yes | 17, 8.5% | 0, 0.0% | 10, 15.4% | 7, 8.2% | 34, 6.2% | Before<br>P=0.111 |
|  |  |  |  |  |  | **After**<br>**P=0.008** |
| **Claustrophobia**<br>Yes | NA |  | 10, 15.2% | 9, 11.1% | NA | NA |
| **Hood Too Loose**<br>Yes | NA |  | 0, 0.0% | 3, 3.6% | NA | NA |
| **Hood Too Tight**<br>Yes | NA |  | 12, 18.2% | 19, 22.6% | NA | NA |
| **Outcomes (n, %)**<br>Died<br>Improved<br>Intubated<br>Unknown | 58, 22.5%<br>200, 77.5%<br>0, 0.0% | 21, 25.9%<br>**57**, 81.4%<br>3, 3.7% | 4, 5.7%<br>**57**, 81.4%<br>9, 12.9% | 41, 46.6%<br>44, 50.0%<br>3, 3.4% | **124, 23%**<br>**358, 65%**<br>**15 3%**<br>**49, 9%** | **Before**<br>**P≤0.001** |
|  |  |  |  |  |  | After<br>P=0.20 |

Consequently, to evaluate outcomes with and without use of the CPAP/O2 helmet, we conducted separate ‘with versus without’ comparisons before and after the training (Table 2). Prior to training, patient outcomes (death, improved, intubated) were significantly better in the group that received CPAP/O2 helmet NIV than those who did not. This difference was not observed after the training. We consequenty stratified patients by their disease severity before treatment, but did not observe without improvement in the assessed outcomes after training.

### Association between CPAP/O2 helmet use and COVID-19 seroprevalence among healthcare workers

We evaluated the association between use of CPAP/O2 helmet NIV and COVID-19 seroprevalence among healthcare workers in order to address the concern that NIV might expose healthcare workers to viral infection [Table 3]. Forty percent of the 282 healthcare workers were seropositive. Of the seropositive subset, 49% were nurses, 25% physicians [Table 4]. There were no statistically significant differences in seroprevalence by days worked on COVID-19 wards, number of patients treated with COVID-19, contact with respiratory secretions or aerosol generating procedures on COVID-19 patients (P>0.10 for all), Additionally, there were no differences in COVID-19 serostatus by the number of CPAP/O2 helmet NIV-treated patients cared for, or number of CPAP/O2 helmet devices or filters cleaned [Table 5].

**Table 3.** Participation of health care workers in the assessment of COVID-19 exposure

|  | Positive anti-body results<br>N=112 |  | Negative anti-body results<br>N=170 |  | All anti-body results<br>N=282 |  |
| --- | --- | --- | --- | --- | --- | --- |
| **COVID-19 Treatment Center** |  |  |  |  |  |  |
| AKTH | 21 | 18.8% | 26 | 15.3% | 47 | 16.7% |
| DELSUTH | 18 | 16.1% | 26 | 15.3% | 44 | 15.6% |
| FMC Lagos | 20 | 17.9% | 37 | 21.8% | 57 | 20.2% |
| LUTH | 9 | 8.0% | 11 | 6.5% | 20 | 7.1% |
| UCH | 25 | 22.3% | 46 | 27.1% | 71 | 25.2% |
| UNTH | 19 | 17.0% | 24 | 14.1% | 43 | 15.2% |
| EUTH | 0 | 0% | 0 | 0% | 0 | 0% |
| FUTH | 0 | 0% | 0 | 0% | 0 | 0% |

**Table 4.**
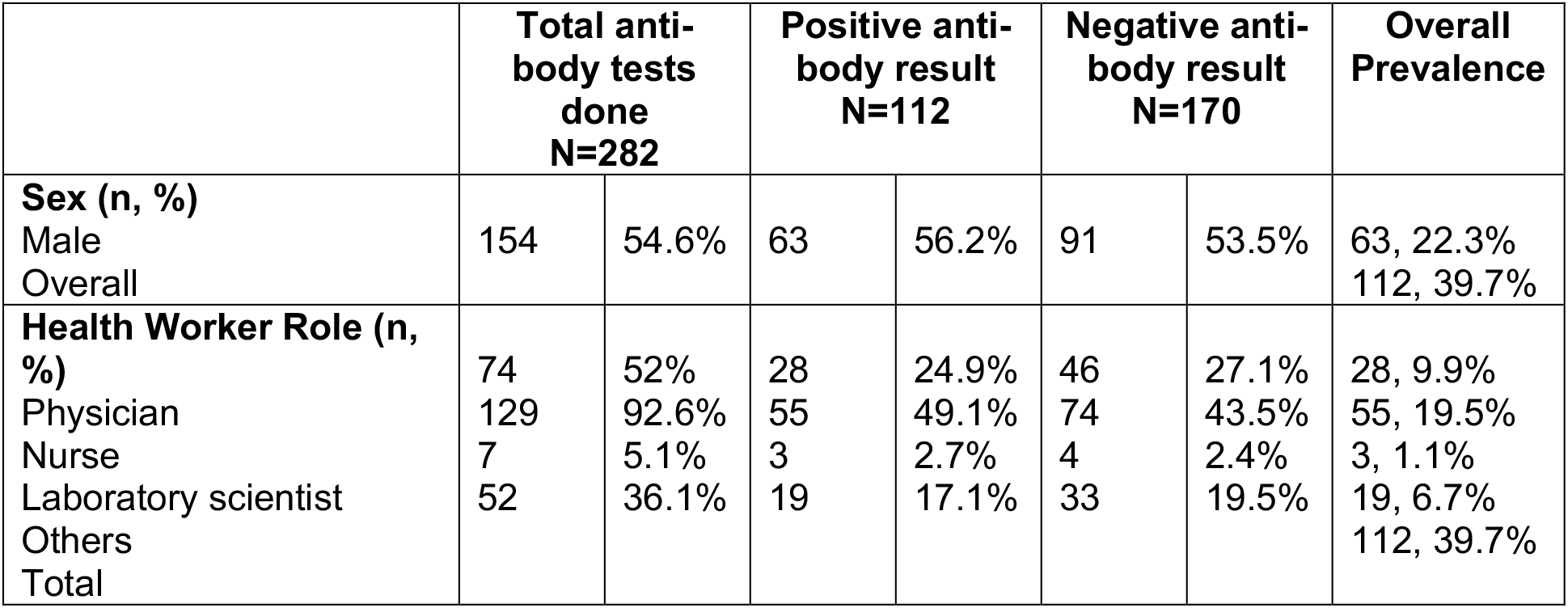
Results of antibody tests in the healthcare worker population.

**Table 5.** Work characteristics of the sample of healthcare workers associated with and without seropositivity for SARS-CoV-2.

|  | **Positive**<br>**anti-body**<br>**result**<br>**N=112** | **Negative**<br>**anti-body**<br>**result**<br>**N=170** | **Total anti-**<br>**body tests**<br>**done**<br>**N=282** | **P-**<br>**Value** |
| --- | --- | --- | --- | --- |
| Days worked on COVID-19 wards<br>Mean ± SD<br>None<br><30 days<br>30-90 days<br>>90 days | 53±94.5<br>44, 39.3%<br>28, 25.0%<br>20, 17.9%<br>20, 17.9% | 73±102.5<br>47, 27.6%<br>43, 25.3%<br>33, 19.4%<br>47, 27.6% | 91, 32.3%<br>71, 25.2%<br>53, 18.8%<br>67, 23.8% | 0.130 |
| COVID-19 helmet patients cared<br>for<br>Mean ± SD<br>None<br>1-2<br>3-4<br>$\geq 5$ | 5.2 ± 10.2<br>54, 48.2%<br>16, 14.3%<br>15, 13.4%<br>27, 24.1% | 7.03±20.1<br>68, 40.0%<br>33, 19.4%<br>29, 17.1%<br>40, 23.5% | 122, 43.3%<br>49, 17.4%<br>44, 15.6%<br>67, 23.8% | 0.446 |
| COVID-19 patients cared for<br>Mean ± SD<br>None<br>< 10<br>10-19<br>20-29<br>$\geq 30$ | 22.6 ± 31.8<br>46, 41.1%<br>14, 12.5%<br>8, 7.1%<br>12, 10.7%<br>32, 28.6% | 26.8±35.6<br>52, 30.6%<br>27, 15.9%<br>19, 11.2%<br>17, 10.0%<br>55, 32.4% | 98, 34.8%<br>41, 14.5%<br>27, 9.6%<br>29, 10.3%<br>87, 30.9% | 0.385 |
| COVID-19 patients whose<br>respiratory secretions were<br>contacted<br>Mean ± SD<br>None<br>1<br>2<br>>2 | 1.7 ± 4.8<br>87, 77.7%<br>4, 3.6%<br>7, 6.3%<br>14, 12.5% | 1.8±4.5<br>122, 71.8%<br>9, 5.3%<br>12, 7.1%<br>27, 15.9% | 209, 74.1%<br>13, 4.6%<br>19, 6.7%<br>41, 14.6% | 0.717 |
| CPAP helmet cleaned<br>Mean ± SD<br>None<br>1<br>2<br>>2 | 0.76±1.6<br>81, 72.3%<br>11, 9.8%<br>6, 5.4%<br>14, 12.5% | 0.98±2.2<br>119, 70.0%<br>14, 8.2%<br>13, 7.6%<br>24, 14.1% | 200, 70.9%<br>25, 8.%<br>19, 6.7%<br>38, 13.5% | 0.826 |
| Filters exchanged on CPAP<br>helmet<br>Mean ± SD<br>None<br>1<br>2<br>>2 | 0.6±1.73<br>85, 75.9%<br>12, 10.7%<br>9, 8.0%<br>6, 5.4% | 1.15±3.3<br>126, 74.1%<br>10, 5.9%<br>15, 8.8%<br>19, 11.2% | 211, 74.8%<br>22, 7.8%<br>24, 8.5%<br>25, 8.9% | 0.197 |
| Use of PPE and hand hygiene on<br>COVID-19 patients<br>Yes<br>No | 64, 98.5%<br>1, 1.5% | 109, 96.5%<br>4, 3.5% | 173, 97.2%<br>5, 2.8% | 0.266 |
| Aerosol generating procedures on COVID-19 patients |  |  |  | 0.422 |
| Yes | 53, 81.5% | 94, 86.2% | 147, 84.5% |  |
| No | 12, 18.5% | 15, 13.8% | 27, 15.5% |  |
| PPE and hand hygiene on COVID-19 patients with AGP's |  |  |  | **0.020** |
| Yes | 56, 90.3% | 102, 99.0% | 158, 95.8% |  |
| No | 6, 9.7% | 1, 1.0% | 7, 4.2% |  |

Only inadequate PPE and hand hygiene during aerosol-generating procedures (AGP) on COVID-19 patients was associated with increased SARS-CoV-2 seropositivity (p=0.02). AGPs include tracheal intubation, mechanical ventilation, nebulizer treatment, manipulation of O2 masks, oxygen administration, chest physiotherapy, chest compression, insertion of nasogastric tube, but not administration of CPAP/O2 helmet treatment. The same increased seroprevalence was not observed for inadquqate PPE during non AGPs.

Finally, with respect to the relationship between community exposure in the healthcare workers and COVID-19 infection among the healthcare workers, there was a 6-fold increased seropositivity likelihood in those who answered ‘yes’ to being within 1m of a person with COVID-19 for 15 minutes or longer in the past 14 days, compared to those who answered ‘yes’ to living in the same household as a person with COVID-19 in the past 14 days (Table 6).

**Table 6.** Community exposure to SARS-CoV-2 in the sample of healthcare workers

| Variables | Positive anti-body result<br>N=112 |  | Negative anti-body result<br>N=170 |  | All anti-body results<br>N=282 |  |
| --- | --- | --- | --- | --- | --- | --- |
| **Overall (all participants)** |  |  |  |  |  |  |
| Yes | 6 | 5.40% | 8 | 4.70% | 14 | 5.00% |
| No | 77 | 68.80% | 124 | 72.90% | 201 | 71.30% |
| Uncertain | 29 | 25.9% | 38 | 22.30% | 67 | 23.80% |
| **Within one meter of a person with COVID-19 for 15 minutes or longer in the past 14 days** |  |  |  |  |  |  |
| Yes | 34 | 30.4% | 49 | 28.8% | 83 | 29.4% |
| No | 41 | 36.6% | 70 | 41.2% | 111 | 39.4% |
| Uncertain | 37 | 28.1% | 51 | 30.0% | 88 | 31.2% |
| **Had direct physical contact with a person with COVID-19 in the past 14 days** |  |  |  |  |  |  |
| Yes | 20 | 17.9% | 36 | 21.2% | 56 | 19.9% |
| No | 57 | 50.9% | 85 | 50.0% | 142 | 50.4% |
| Uncertain | 35 | 31.3% | 49 | 28.8% | 84 | 29.7% |

## DISCUSSION

There is growing evidence of the potential benefits of helmet-based CPAP NIV for the management of respiratory failure among patients with COVID-19 (1, 19). Our CircumVent project demonstrates the feasibility and acceptability of training healthcare providers in a LMIC to efficiently deploy this strategy in the course of the COVID-19 pandemic utilizing practice facilitation. Importantly, treatment of patients with CPAP/O2 helmet NIV did not appear to be pose an increased risk of COVID-19 infection to healthcare providers.

Efficienct deployment of CPAP/O2 helmet NIV was aided by the familiarity that most providers in the CircumVent network had with CPAP NIV. Though the helmet device itself was novel, providers felt that it was easy to use. Accordingly, we observed appropriate use of the helmet on patients with more comorbid conditions, and more severe disease as evidenced by more severe tachypnea and hypoxia even before training through practice facilitation. However, we still observed benefits from practice facilitation, a goal-oriented, context-dependent social process for implementing new knowledge into practice or organizational routines. At the provider level, we observerd a greater prevalence of comorbid conditions among CPAP/O2 helmet NIV-treated patients after facilitation, a finding which may have represented more concerted selection of higher risk patients. Indeed the evidence is clear that patients with certain chronic cardiovascular, pulmonary, metabolic, and immunologic conditions have a higher risk of progressive to severe COVID-19 disease, and these patients would be the most likely to need respiratory support. In addition to the benefits for provider practice, the process of practice facilitation allowed us to rapidly identify and address implementation barriers including minimizing oxygen waste, troublehooting back-up power options, and sourcing consumables for the helmet circuit. This process can be replicated in other environemnts which may have additional, context-specific barriers to address during deployment and scale-up of this treatment protocol.

Clinical effectiveness was outside of the scope of this implementation study, but the mortality observed in our cohort overall (23%), and among patients treated with CPAP/O2 helmet NIV was consistent with that observed in similar settings in Sub-Saharan Africa. Particularly in the context of adopting a new treatment strategy, it was important that the strategy not pose any additional risk of infection to healthcare workers beyond their standard of care. The data on risk of transmission of COVID-19 and indeed other respitory viruses from AGPs has been inconsistent. Though the threat of COVID-19 infection to healthcare workers may be less in in the era of COVID-19 vaccination, widespread vaccination remains the exception in LMIC, and evolving variants of SARS-CoV-2 have shown increased transmissibility, thus weakening vaccine-mediated protection from infection that we witnessed earlier in the pandemic. Notably, our seroprevalence study demonstrated no evidence of increased risk of infection among healthcare workers who treated patients with the CPAP/O2 helmet NIV, or who managed or cleaned the helmet devices. Like other healthcare worker seroprevalence studies in the region, the seroprevalence in our cohort was quite high (40%), suggesting the possibility of both high community and hospital transmission, but also possibly limiting our power to assess the relationship between exposures and risk. Interestingly, our seroprevalence study only identified improper PPE and hand hygiene in the context of an AGP as a risk factor for seropositivity, providing some reassurance that the level of risk for those procedures might be higher than for managing the CPAP/O2 helmet.

Despite the important implications of our findings, our study has some weaknesses. The study was primarily motivated to supply a low-cost strategy for respiratory support during the COVID-19 pandemic, an effort untertaken by volunteers. These data were collected at convenience within the context of that implementation, and consequently are limited compared to, for example, a randomized intervention-control, two-arm implementation study designed primarily to assess implementation and/or clinical goals. Nonetheless, the findings are clear and potentially impactful. CPAP/O2 helmet NIV can be implemented by healthcare workers in a LMIC with the protocol (see www.circumventproject.com/protocol) and minimal formal training. The implementation can effectively improve COVID-19 patient outcomes, including to reduce mortality, all with no evidence of harm to the healthcare workers from increased risk of infection.

## CONCLUSIONS

The evolution of COVID-19 B.1.617.2 (Delta variant), identified in October 2020, left India devastated by a third wave of infection with daily confirmed cases peaking at over 2 million, before becoming the predominant strain worldwide (20). Similarly, COVID-19 B.1.1.529 (Omicron variant) was first identified in South Africa and Botswana in November 2021 before surging across the globe (21). Possible widespread circulation of more devastating strains represent the most pressing recent example of the ongoing need to provide innovative care delivery solutions for managing COVID-19 (8-10, 21). Deploying cost-effective treatment strategies built on this model could be considered preemptively and during crisis mitigation phases to increase the capacity for providers to manage severe respiratory infections in regions with limited infrastructure for invasive mechanical ventilation. Further, this approach can also build capacity of LMIC health facilities to manage respiratory failure from common respiratory pathogens.

## Data Availability

All data produced in the present study are available upon reasonable request to the authors

## ACKNOWLEDGEMENTS

Funding support by the CDC Foundation award number 1085.1 and the Aliko Dangote Foundation grant number GS2008-0001. We are grateful to the 40+ volunteers who worked to develop the concept of the CPAP/O2 Helmet system within the Ventilator Project (www.ventilatorproject.org), especially volunteer John Gridley who guided and helped the team to navigate the medical device and funding spaces, and volunteer Deepika Grover who helped with communications and the search for funding. We would like to thank the non-profit group Ocean Opportunity for their generous donation of expertise and materials used in this work. We are grateful to Michael Nottidge MD, MPH, MBA FACEP for his expert guidance in protocol development and provider training, Sani Aliyu, MBBS, FRCP, National Coordinator of the Nigerian Presidential Task Force of COVID-19, and the Federal Ministry of Health, Nigeria for their support.

## AUTHOR CONTRIBUTIONS

Conceptualization: AAA, ABS, AAF, AH, BLS, GO, IO, MHA Data

Curation: AZM

Formal Analysis: AZM, AAF, AH

Funding Acquisition: AAF, BLS, GO

Investigation: AA, AAO, OA, NAA, SA, HA, EBA, ACA, AAD, COF, IO, II, SM, AAM, BMM, IIU, KNU, USU, OAU, CU, MKR, IEA, NIN, OO, TOO, AIS, WLA

Methodology: AAA, AAF, AH, GO, IO, MHA

Project Administration: AAF, BLS Resources: AH

Supervision: AAF, BLS, OCE

Writing: AAA, AAF, GO

